# Impact of the COVID-19 pandemic on infant and pediatric asthma: a multi-center survey using an administrative database in Japan

**DOI:** 10.1101/2020.11.29.20240374

**Authors:** Seiko Bun, Kenji Kishimoto, Jung-ho Shin, Daisuke Takada, Tetsuji Morishita, Susumu Kunisawa, Yuichi Imanaka

**Author notes:** Correspondence: Yuichi Imanaka, MD, Ph.D, Department of Healthcare Economics and Quality Management, Graduate School of Medicine, Kyoto University, Yoshida Konoe-cho, Sakyo-ku, Kyoto 606-8501, JAPAN. Alternate corresponding author: Seiko Bun, Department of Healthcare Economics and Quality Management, Graduate School of Medicine, Kyoto University, Yoshida Konoe-cho, Kyoto 606-8501, Japan.

## Abstract

**Background:** Limited data are available on the relationship between infant and pediatric asthma and severe acute respiratory syndrome coronavirus 2 (COVID-19).

Political limitations such as school closure may affect the treatment behavior of pediatric asthma. To investigate the trends of treatment behavior in the field of pediatrics during the COVID-19 pandemic.

**Methods:** This is a retrospective observational study using Diagnosis Procedure Combination (DPC) data from the Quality Indicator/Improvement Project (QIP) database. We identified children with asthma aged 15 years or younger who were patients from July 1, 2018, to June 30, 2020. The main outcome was a comparison between asthma patients’ treatment behavior before the COVID-19 pandemic and during the COVID-19 pandemic.

We statistically tested the admission volume changes based on the discharge date after adjusting for seasonality through a Fourier term using an interrupted time-series analysis (ITS).

**Results:** We identified 10,481 inpatients cases in 67 hospitals and 258,911 out-patients cases in 180 hospitals who were diagnosed with asthma. We performed ITS analysis for inpatients. The reduction in the number of patients during this period was estimated to be 232 (*P=*0.001). In addition, ITS analysis was performed for patients aged <3 years. The reduced number of patients during this period was estimated to be 155 (P<0.001).

**Conclusions:** We found that the number of pediatric asthma patients dramatically decreased during the COVID-19 pandemic. We need to continue research into the trends of pediatric asthma patients after the COVID-19 pandemic in Japan.

**Key Messages:** There are increasingly available data on the relationship between adults’ asthma and COVID-19. However, in the fields of pediatrics, limited data are available. Patients with moderate to severe asthma who needed hospitalization dramatically decreased during the COVID-19 pandemic. Besides, doctors prescribed nebulizers more than metered-dose inhalers by the clinical guideline recommendation. Our findings reinforce the value of political inventions, such as school closure reduced the number of asthma attacks in infants and school-age children.

## Introduction

In December 2019, pneumonia caused by a novel coronavirus (SARS-CoV-2) occurred in Wuhan, China. According to the World Health Organization (WHO), 217,769 deaths had been caused by coronavirus disease 2019 (COVID-19) globally by May 1, 2020^1^. Public health interventions, including school closure and the city lockdown, began in many countries.

Pediatric asthma is one of the most common diseases in infant to school-age children. Risk factors, including respiratory virus infection, air pollution, and house dust, can exacerbate pediatric asthma. Environmental triggers are particularly important. In Japan, all schools excluding nursery schools were closed nationwide on February 27, 2020^2^. Besides, the COVID-19 pandemic caused dramatic changes in daily practice and health care delivery systems in many countries. Some states reported hospital avoidance in pediatrics, including emergency department visits and hospital admission during the COVID-19 pandemic.

Unlike the lockdown policy in many countries, including the United States, China, and European countries, the Japanese lockdown did not have a legally binding force. Therefore, the lockdown in Japan was a voluntary lockdown^2^. The ‘voluntary lockdown’ promoted a change of lifestyle, such as decreasing transportation and industrial activity and closure of schools, which intended to limit physical interactions among people. These political interventions may have affected the environment for children.

In this study, we describe the impact of the COVID-19 pandemic on pediatric asthma-related health care utilization including emergency departments and hospitalization.

Inhaled steroids and β_2_ agonists are a key treatment option for asthma^3^. The Japanese Society of Pediatric Allergy and Clinical Immunology recommends using a metered-dose inhaler rather than a nebulizer because a nebulizer can spread the aerosol on caregivers and medical staff. We also investigated the trends for the prescription of inhalers^4^.

## Methods

### Source of data

Our study was a retrospective observational study using Diagnosis Procedure Combination (DPC) data from the Quality Indicator/Improvement Project (QIP) database. The QIP database contains DPC data from acute care hospitals voluntarily participating in the project. The DPC/per-diem payment system (PDPS) is a Japanese prospective payment system applied to acute care hospitals. In total, 1,730 hospitals adopted the DPC/PDPS in 2018, which accounted for 54% (482,618/891,872) of general beds of Japanese hospitals in 2018^5,6^

The cumulative number of participating hospitals for the QIP was over 500, which were located all over Japan and included both public and private hospitals of various sizes. The number of general beds, which are hospital beds that are not psychiatric, infectious diseases, or tuberculosis beds according to the Japanese classification of hospital beds, ranged from 30 to 1,151 in 2016. The DPC data consist of claims and discharge summaries, including International Classification of Diseases, 10^th^ Revision (ICD-10) codes classifying the primary diagnosis, cause of admission, the most and the second-most medical-resource-intensive diagnoses, up to 10 comorbidities, and 10 complications. The DPC data also contain codes of all services and medications provided during each hospitalization as well as PDPS information.

### Study population

The study was conducted after obtaining approval from the Kyoto University Hospital Ethics committee of the Kyoto University Graduate School and Faculty of Medicine, according to the guidelines on medical and epidemiological research (No. R0135). Using the DPC database, we identified inpatients with discharge dates from July 1, 2018 to June 30, 2020, and outpatients at the date of a visiting hospital within the same periods.

We collected data for each patient during the research period. We selected hospitals that sent DPC data for more than 9 months each year to QIP database.

All patients were aged ≤15 years and were diagnosed with asthma according to the ICD-10 codes (J45.x, J46).

### Statistical analysis

An interrupted time-series analysis (ITS), including segmented regressions, was performed to evaluate the effect of the COVID-19 pandemic on population-level admissions. We statistically tested the changes in admission volume based on the discharge date after adjusting for seasonality through a Fourier term. We compared the number of admissions per month based on the discharge date, not the date of admission, because the DPC data were generated after patients’ discharge from hospitals. We hypothesize that COVID-19 would impact the level change of admission volume immediately after March 2020, as an implemented point. The Japanese government declared a state of emergency and requested self-quarantine, social distancing, and school closure. We classified the patients into three age groups: 0–2, 3–5, and 6–15 years. These groups corresponded to nursery school for ages 0–2, kindergarten for ages 3–5 years, elementary school for ages 6–12 years, and junior high school for ages 12–15 years.

All analyses were computed using R statistical software (version 3.4.0), and a two-sided significance level was fixed at 0.05.

## Results

### Studied patients

Table 1 shows the patients’ characteristics. We identified 10,481 in-patient cases in 67 hospitals and 258,911 out-patient cases from 180 hospitals, who were diagnosed with asthma. The median age was 2 years (IQR 1, 5 years), and the most common age category was 0–2 years in inpatients. The median age was 5 years (IQR 3, 9 years), and the most common age category was 6–11 years in outpatients. The number of males was much bigger than females in both inpatients and outpatients. The median value of the length of stay for each month was 5 days (IQR 4, 6) during the study period. The proportion of patients with asthma who visited emergency departments from July 2019 to June 2020 decreased by more than 30% compared to that from July 2018 to June 2019.

**Table 1.** Patients characteristics n-patient characteristics.

|  | Total period | July 1, 2018 to June 30, 2019 | July 1, 2018 to June 30, 2020 |
| --- | --- | --- | --- |
| Number of patients | 10,481 | 6,245 | 4,236 |
| (Special vigilance area) | (5,929) | (3,473) | (2,456) |
| Infectious disease medical institution | 28 | 28 | 28 |
| Median age (IQR) | 2 (1, 5) |  |  |
| 0–2 | 4,307 | 2,698 | 1,825 |
| 3–5 | 2,563 | 1,574 | 1,110 |
| 6–11 | 1,582 | 999 | 667 |
| 12–15 | 241 | 142 | 111 |
| Sex |  |  |  |
| Male | 5,190 |  |  |
| Female | 3,354 |  |  |
| Average length of stay (IQR) | 5 (4, 6) |  |  |
| Urgency admission (ambulance) | 10,287 (937) | 6,144 (523) | 4,143 (414) |
| Intensive care unit admission | 39 | 18 | 21 |

**b) Out-patient characteristics**
|  | Total period | July 1, 2018 to June 30, 2019 | July 1, 2018 to June 30, 2020 |
| --- | --- | --- | --- |
| Number of visited patients | 258,911 | 147,321 | 111,590 |
| Hospitals in special vigilance area | 98 | 94 | 91 |
| Infectious disease medical institution | 42 | 42 | 41 |
| Median age (IQR) | 5 (3, 9) |  |  |
| 0–2 | 12,664 | 8,766 | 5,988 |
| 3–5 | 12,934 | 9,016 | 7,061 |
| 6–11 | 14,452 | 10,751 | 8,806 |
| 12–15 | 5,305 | 3,903 | 3,260 |
| Sex |  |  |  |
| Male | 23,922 |  |  |
| Female | 16,476 |  |  |

### Asthma patients during the COVID-19 pandemic

Figure 1 shows the number of inpatients between July 2019 and June 2020. There was a seasonal increase from March to June 2019. However, the number of inpatients from March to June 2020 decreased (Figure 1 a)). Figure 1 b) shows the results of the interrupted time-series analysis. The results showed a marked reduction in the number of inpatients from March 2020 to June 2020; the reduced number of patients during this period was estimated to be 232 (*P=*0.0012). Among the analyses for each age category, the number of inpatients aged ≤3 years between July 2019 and June 2020 showed a statistically significant decrease compared to that between July 2018 and June 2019. The number of inpatients of the other age categories also showed decreasing trends between July 2019 and June 2020 compared to that between July 2018 and June 2019 (Figure 2 a)–c)).

**Figure 1.**
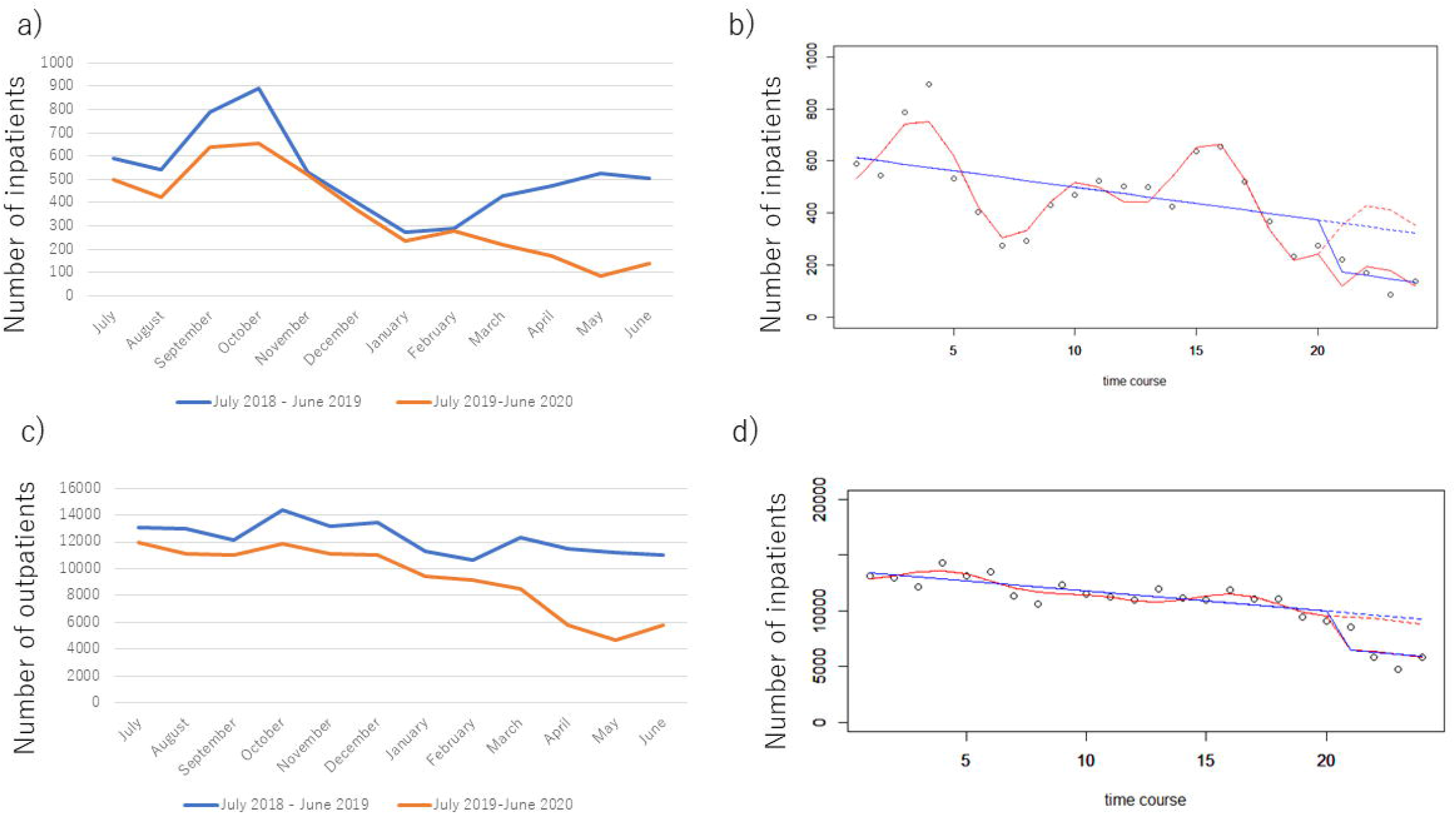
Asthma patients during the COVID-19 pandemic. a) Trends of cases of inpatients between July 2018 to June 2019 and July 2019 to June 2020. b) An interrupted time-series (ITS) analysis for inpatients. The reduced number of patients during this period was estimated to be 232 (*P=*0.0012). c) Trends of cases of outpatients between July 2018 to June 2019 and July 2019 to June 2020. d) ITS analysis for outpatients. The reduced number of patients during this period was estimated to be 2,975 (*P=*0.0010).

**Figure 2.**
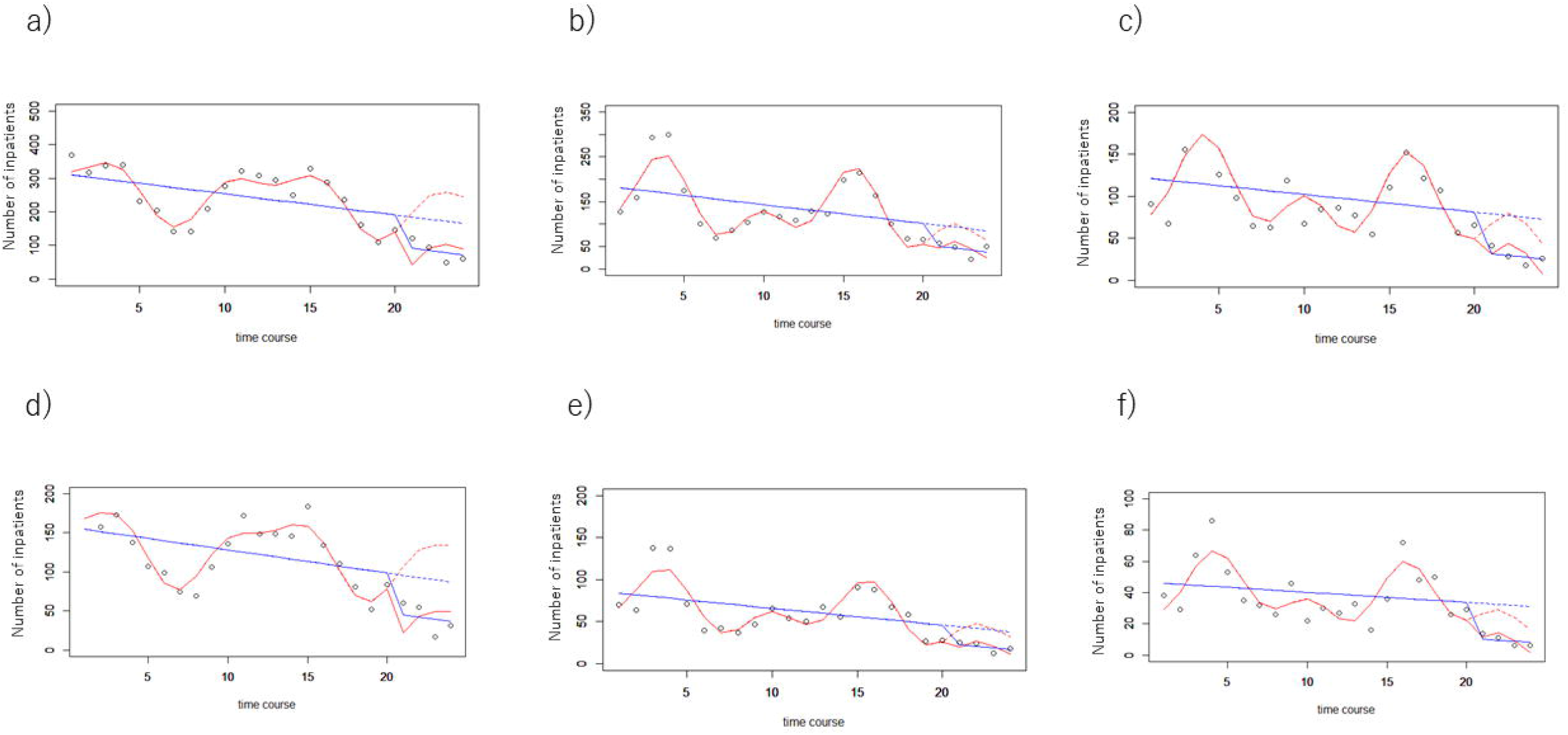
Asthma inpatients during the COVID-19 pandemic for each age category. a) An interrupted time-series (ITS) analysis for patients aged <3 years. The reduced number of patients during this period was estimated to be 155 (*P*<0.001). b) ITS analysis for patients aged 3–5 years. The reduced number of patients during this period was estimated to be 20 (*P=*0.118). c) ITS analysis for patients aged ≥6 years. The reduced number of patients during this period was estimated to be 36 (*P=*0.164). d) ITS analysis for patients aged <3 years who were complicated with an infection. The reduced number of patients during this period was estimated to be 84 (*P*<0.001). e) ITS analysis for patients aged 3–5 years who were complicated with an infection. The reduced number of patients during this period was estimated to be 39 (*P=*0.086). f) ITS analysis for patients aged ≥6 years who were complicated with an infection. The reduced number of patients during this period was estimated to be 14 (*P=*0.139).

The number of patients who were complicated with an infection showed a statistically significant decrease between July 2019 and June 2020 compared to that between July 2018 and June 2019, especially for patients aged ≤3 years (Figure 2 d)–f)). There was a statistically significant decrease in the number of outpatient cases between July 2018 to June 2019 and July 2019 to June 2020 (Figure 1c), d)).

### Trends for the prescription of inhalers

The prescription number of both inhalers and nebulizers for inpatients decreased during the COVID-19 pandemic. Also, the proportion of nebulizers decreased by more than that of metered-dose inhalers. (Figure 3)

**Figure 3.**
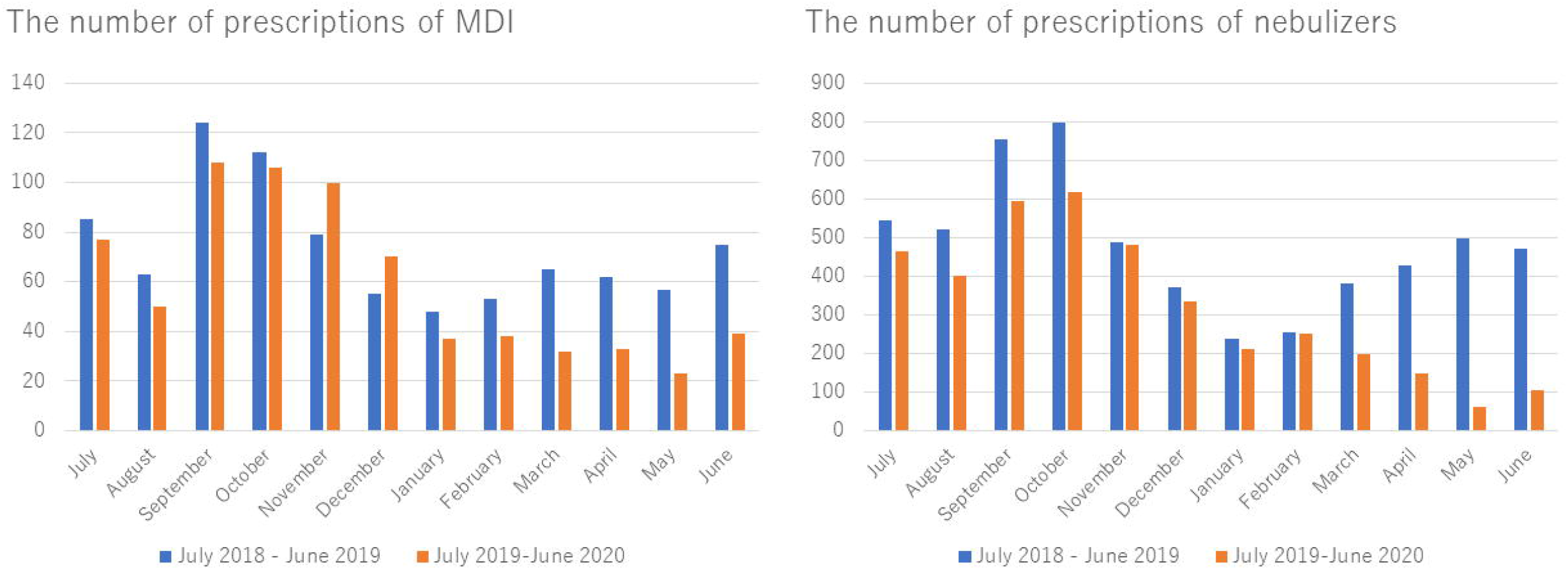
Trends for the prescription of inhalers. The proportion of nebulizers decreased by more than that of metered-dose inhalers.

## Discussion

This study demonstrated the trends of asthma in the pediatric population by using a large-scale administrative database. Our study had three major findings.

First, pediatric patients with moderate to severe asthma who needed hospitalization dramatically decreased during the COVID-19 pandemic. Moreover, infant patients controlled their asthma attacks more than school-age children. There were several reasons for this situation. One reason for asthma is an infection^7^; school closure and ‘voluntary lockdown’ may have influenced the children’s disease. The most common route of infection is contact among children and is well known. Besides, the Japanese government convinced businessmen to stay and work at home during the ‘voluntary lockdown’ starting in April ^8^. Previous reports have shown that adherence improved during lockdown for the COVID-19 pandemic^9^. Besides, PM2.5 was one of the environmental risk factors that decreased in several areas, including India, China, England, and The United States during lockdown. Lockdown caused a reduction in traffic volume, including cars, airplanes, and trains, which resulted in decreased environmental risk factors, including PM2.5, CO_2_, and O_3_^10,11,12^.

Second, the number of outpatients during the pandemic decreased compared to that before the pandemic. Most outpatients controlled their asthma, so it is possible that the patient’s parents avoided a hospital visit. These hospital avoidances in Japan were similar to other countries^13^. Previous reports have stated that online or telephone health care services increased in European countries^14^. Online or telephone health care services could have been provided in Japan; however, there were few cases using online or telephone health care services in our study (data not shown).

Third, doctors prescribed nebulizers more than metered-dose inhalers. The Japanese Society of Pediatric Allergy and Clinical immunology recommended using a metered-dose inhaler rather than a nebulizer in March 2020, because a nebulizer can spread aerosol on caregivers and medical staff. The guideline has not infiltrated sufficiently to all hospitals and medical staff during a short period, and infant patients cannot use a metered-dose inhaler.

Our study had several limitations. QIP hospitals represented a limited number of hospitals in Japan. Therefore, it is difficult to generalize our results to nationwide pediatric asthma patients. However, a previous study^15^ for adult’s asthma patients showed similar results to ours. Therefore, we consider that our findings also have validity for inpatients. Although most outpatients with asthma usually visit clinics, acute hospitals such as QIP hospitals treat patients who have other comorbidities or follow the acute phase for asthma. Finally, we had no data about air pollution in Japan.

## Conclusions

We found that the number of pediatric asthma patients did not increase due to respiratory infections such as COVID-19. These results were supported by political inventions such as school closure and social distancing. In addition, infant patients controlled their asthma attacks more than school-age children. We need to continue the research into trends of pediatric asthma patients after the COVID-19 pandemic in Japan.

## Data Availability

The datasets generated during and/or analyzed during the current study are available from the corresponding author on reasonable request.

## Acknowledgments

This study was supported by JSPS KAKENHI Grant Number JP19H01075 from the Japan Society for the Promotion of Science, Health Labour Sciences Research Grant from the Ministry of Health, Labour and Welfare, Japan [20HA2003] and by the GAP Fund Program of Kyoto University type B to Yuichi Imanaka. The funders played no role in the study design, data collection and analysis, decision to publish, or preparation of the manuscript.

